# Genome-wide association study identifies two new loci associated with anti-NMDAR encephalitis

**DOI:** 10.1101/2021.03.11.21253347

**Authors:** Anja K Tietz, Klemens Angstwurm, Tobias Baumgartner, Kathrin Doppler, Katharina Eisenhut, Martin Elišák, Andre Franke, Kristin S Golombeck, Robert Handreka, Max Kaufmann, Markus Kraemer, Andrea Kraft, Jan Lewerenz, Wolfgang Lieb, Marie Madlener, Nico Melzer, Hana Mojzisova, Peter Möller, Thomas Pfefferkorn, Harald Prüss, Kevin Rostásy, Margret Schnegelsberg, Ina Schröder, Kai Siebenbrodt, Kurt-Wolfram Sühs, Klaus-Peter Wandinger, Jonathan Wickel, Frank Leypoldt, Gregor Kuhlenbäumer, on behalf of the German Network for Research on Autoimmune Encephalitis (GENERATE)

**Author notes:** **Corresponding author**: Gregor Kuhlenbäumer, Department of Neurology, University Hospital Schleswig-Holstein Kiel, Arnold-Heller-Str. 3, 24105 Kiel Germany.

## Abstract

**Objective:** To investigate the genetic determinants of the most common type of antibody-mediated autoimmune encephalitis, anti-N-methyl-D-aspartate receptor (anti-NMDAR) encephalitis.

**Methods:** We performed a genome-wide association study in 178 patients with anti-NMDAR encephalitis and 590 healthy controls followed by a colocalization analysis to identify putatively causal genes.

**Results:** We identified two independent risk loci harboring genome-wide significant variants (P < 5 × 10^−8^, OR ≥ 2.2), one on chromosome 15, harboring only the *LRRK1* gene, and one on chromosome 11 centered on the *ACP2* and *NR1H3* genes in a larger region of high linkage-disequilibrium. Colocalization signals with expression quantitative trait loci (eQTL) for different brain regions and immune cell types suggested *ACP2, NR1H3, MADD, DDB2*, and *C11orf49* as putatively causal genes. The best candidate genes in each region are *LRRK1*, encoding Leucine-Rich Repeat Kinase 1, a protein involved in B-cell development, and *NR1H3* liver x receptor alpha, a transcription factor whose activation inhibits inflammatory processes.

**Conclusion:** This study provides evidence for relevant genetic determinants of antibody-mediated autoimmune encephalitides outside the HLA-region. The results suggest that future studies with larger sample sizes will successfully identify additional genetic determinants and contribute to the elucidation of the pathomechanism.

## Introduction

Antibody-mediated encephalitides are a recently discovered group of rare diseases caused by autoantibodies against central nervous system (CNS) system antigens.^1^ Subgroups are defined by the respective target antigens. The most common subgroup is caused by immunoglobulin-G (IgG) class antibodies against the GluN1 subunit of the N-methyl-D-aspartate (NMDA) receptor (NMDAR), the most important excitatory neurotransmitter in the CNS. NMDAR antibodies cause internalization of surface NMDAR, thereby reducing signal transduction. Anti-NMDAR encephalitis affects children and adults with a female preponderance. The estimated prevalence is 0.6/100,000 population.^2^ It manifests with behavioral changes, psychiatric symptoms, epileptic seizures, memory dysfunction, movement disorders, and loss of consciousness and often responds favorably to immunotherapy.^3^ A definite diagnosis requires the detection of anti-NMDAR antibodies of the IgG class in serum and/or cerebrospinal fluid (CSF). Known trigger factors include ovarian teratomas with ectopic expression of NMDAR and viral (mostly HSV-1) encephalitis.^1^ In a first genome-wide association study (GWAS) of antibody-mediated encephalitides in a much smaller sample, we found no variants showing genome-wide significant association with anti-NMDAR encephalitis.^4^ For this study, we doubled the sample size, modified the analysis parameters, and added colocalization analysis to identify putatively causal genes.

## Materials and Methods

### Study population

In this case-control study, we analyzed 212 samples from patients with NMDAR antibodies (197 German and 15 Czech patients) collected in the years 2015-2020. Anti-NMDAR encephalitis was classified according to consensus criteria based on a compatible clinical syndrome together with detection of IgG-NMDAR antibodies in serum and/or CSF.^3^ One hundred and seven patients were already included in our previous GWAS.^4^ The additional 105 individuals were either recruited via the German Network for Research on Autoimmune Encephalitis (GENERATE, n = 80) or specifically for this study (n = 25). For recruitment, we contacted the centers of the GENERATE network as well other neurological departments caring for patients with antibody-mediated encephalitides. All contributing scientists are listed in **Supplementary Table S1**. Healthy control samples (n = 1,219) were obtained from the PopGen study, a population-based biobank from northern Germany.^5, 6^

### Genotyping

Genomic DNA was isolated from blood (n = 150) or saliva (n = 62) using standard procedures. The samples were genotyped in 3 batches at the Institute of Clinical Molecular Biology, Kiel University on Infinium Global Screening Array (GSA; Illumina, USA). Array version 2.0 was used for cases and version 1.0 for healthy controls. Genotypes were called using Illumina GenomeStudio 2.0 according to the manufacturer’s instructions using in-house cluster-files. We previously determined a >99.8% genotype concordance for DNA isolated from blood and saliva genotyped on the GSA array in eight individuals.

### Quality control and imputation

We used PLINK v.1.9,^7^ R v.3.6.3,^8^ and the Illumina GenomeStudio for genotype quality control. First, we excluded all non-overlapping variants between the two different versions of the GSA-chip, variants with multi-character allele codes, insertions, deletions, duplicated markers, and ambiguous A/T and G/C variants. We determined genotyping sex by the X-chromosome inbreeding coefficients with F < 0.2 being female and F > 0.8 being male and excluded samples with discordance between reported and imputed sex. After that, we filtered first variants then individuals with a relaxed threshold for a call rate of less than 85% followed by a stringent threshold of 98%. We applied a minor allele frequency (MAF) filter of 1%, as well as filters for significant deviation from Hardy-Weinberg equilibrium (HWE; P < 1 × 10^−6^) in controls, informative missingness (P < 1 × 10^−5^) and outlying heterozygosity rate (Mean ± 3 standard deviations). To determine duplicated or cryptically related individuals, we used pairwise genome-wide estimates of the proportion of identity by descent (IBD) on a pruned dataset containing only markers in low LD regions (pairwise r^2^ < 0.2) and excluded those more closely related than third-degree relatives (IBD > 0.125). Of each identified sample pair, we kept the individual with ahigher call rate. To identify ethnic outliers, we used a procedure similar to the one suggested in the R package plinkQC^9^: We combined the genotype data with the samples of the publically available 1000 Genomes Project^10^ and performed a principal component (PC) analysis on the merged dataset. A European center was determined by the first two PCs of known European samples and the Euclidean distance from this center determined the ethnical assignment with samples more than 1.5 times the maximal European Euclidean distance away from the center being excluded. The remaining individuals were utilized for preliminary association analysis based on which we visually inspected the cluster plots of all variants with a P-value < 10^−4^ and discarded variants without adequate cluster separation. To overcome issues with population stratification we matched controls by ancestry and sex to cases with the R package PCAmatchR,^11^ leading to 590 control samples for the analysis and approximately 3 controls per case. An exact match on sex was employed since there were significantly more female samples in the case samples than in the control samples.

Imputation was carried out on the quality assured dataset, containing 768 individuals (590 controls, 178 cases) and 446,353variants. Subsequently, 26,356,529 variants were imputed based on the TOPMed r2 panel^12^ using the TOPMed Imputation Server^13^ which employs Minimac4 for imputation.^14^ A quality check was carried out, including variants with a MAF > 1%, an imputation quality score R^2^ > 0.7, and no significant deviation from HWE (P < 1 × 10^−6^) in controls, resulting in 8,073,349 variants.

### Association Analysis

We conducted an association analysis on the whole dataset employing a genome-wide significance threshold of P < 5 × 10^−8^. We applied an additive logistic regression model, including sex and PCs, to estimate the association of each SNP with the disease status. The number of PCs was chosen using scree plot analysis.^15^ Population stratification was examined using the inflation factor λ and the visual inspection of quantile-quantile (QQ) plots. To further distinguish between confounding factors like population stratification and polygenicity of the anti-NMDAR encephalitis trait, we performed LD score regression (LDSC) using the LDHub web interface.^16^ Conditional analyses in which successively each genome-wide significant variant was included as a covariate were conducted to identify adjacent independent signals. We used 7,122 genotyped and quality controlled variants from the human Major Histocompatibility Complex region on chromosome 6 to impute four-digit Human Leukocyte Antigen (HLA) alleles using the R package HIBAG^17^. It employs attribute bagging to impute genotypes and we chose a prediction model specifically for European ancestry and the Illumina GSA-chip. We performed the association analysis with PyHLA^18^ using an additive, logistic model including sex and the first PC as covariates and adjusted P-values with the Benjamini-Hochberg false discovery rate step-up method.

To examine the origin of the variant-trait association signals more closely we analyzed the subsamples of patients with early/late disease onset (< or ≥ 25 years) and patients with/without a tumor.

To prioritize genes putatively involved in the disease etiology, we investigated the overlap of eQTL from the Genotype-Tissue Expression (GTEx) project^19^ as well as immune cell eQTL from the Blueprint project^20^ and variants in the risk loci identified by this GWAS. We investigated whether these two independent signals might stem from the same causal variant by colocalization analysis conducted with coloc.^21^ Coloc utilizes approximate Bayes factors to estimate posterior probabilities for common variants causal in the GWAS as well as the eQTL study. We studied all variants present in the GWAS results as well as in GTEx V7 for the 13 available brain tissues or present in the Blueprint immune cell eQTL data within 100 kb up-and downstream of each gene in the two encephalitis risk loci. Coloc estimates posterior probabilities (PP) for four different scenarios. PP4 denotes the posterior probability that both traits – the disease association and the eQTL – are caused by the same variant. A PP4 over 70% was considered as evidence for colocalization. We used LocusZoom^22^ and R to visualize the association results. All analyses and the presentation of the results in this manuscript are based on the human genome version 38 (GRCh38/hg38).

### Standard Protocol Approvals, Registrations, and Patient Consents

All participants gave written informed consent. Institutional review board approval was obtained from the ethical advisory boards of the Universities of Kiel and Luebeck (B337/13;13-162).

## Results

**Table 1** summarizes the clinical features of patients and control individuals demonstrating that patients included in our first GWAS^4^ are comparable to the additional patients in this study regarding age, sex, and clinical features. However, the control individuals were much older than the patients. Genotype data for 212 individuals with anti-NMDAR encephalitis and 1,219 controls were available for analysis. After quality control procedures and control matching, 178 cases and 590 healthy controls remained (**Supplementary Table S2**). Imputation resulted in 8,073,349 quality-assured variants with a MAF > 1%. We incorporated sex and the first dimension of the PC analysis as indicated by scree plot analysis as covariates. In contrast to our first GWAS of anti-NMDAR encephalitis, we did not include age as a covariate (for rationale see: **Discussion**). The genomic inflation factor of λ = 1.03 indicated a low degree of population stratification (**Figure 1A**). The LDSC intercept was 1.01 (standard error = 0.01) with 44% of the genomic inflation attributable to confounding bias including population stratification and cryptic relatedness. This indicates that the majority of inflation is caused by polygenicity. We found 13 genetic variants in two distinct loci below the threshold of P = 5 × 10^−8^ for genome-wide significance (**Figure 1B-D, Table 2**) with leading variants rs10902588 on chromosome 15 (odds ratio (OR) = 2.24, [95% confidence interval (CI) = 1.70-2.95], P = 1.78 × 10^−8^) and rs75393320 on chromosome 11 (OR = 2.20 [1.66-2.92], P = 3.78 × 10^−8^) as well as only 14 Kb further downstream rs11039155 with the same P-value and OR. Conditional analysis including the top variants at each locus argues against the presence of any independent secondary signals (**Supplementary Figure S1**). The significant variants on chromosome 15 are located in the *LRRK1* gene encoding a leucine-rich repeat kinase. In the chromosome 11 locus, rs75393320 lies in the Lysosomal Acid Phosphatase 2 (*ACP2*) gene and rs11039155 is located in the Nuclear Receptor Subfamily 1 Group H Member 3 (*NR1H3*) gene.

**Table 1.**
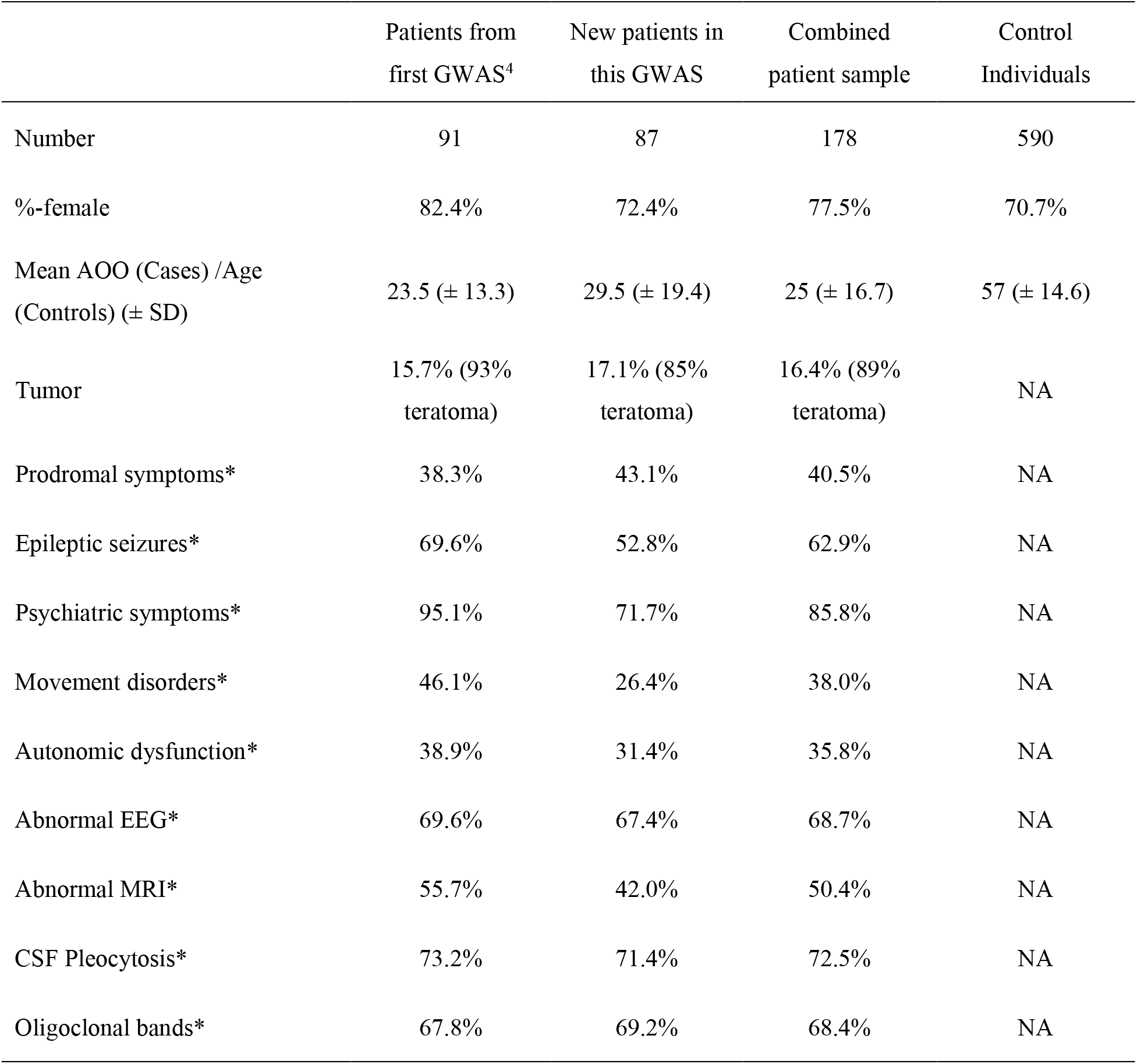
Sample characteristics. Descriptive statistics for the overall patient sample, for patients from the first GWAS^4^, for newly recruited patients and for healthy control individuals. AOO = age of onset, SD = standard deviation, NA = not applicable, *=only available for GENERATE samples.

**Figure 1.**
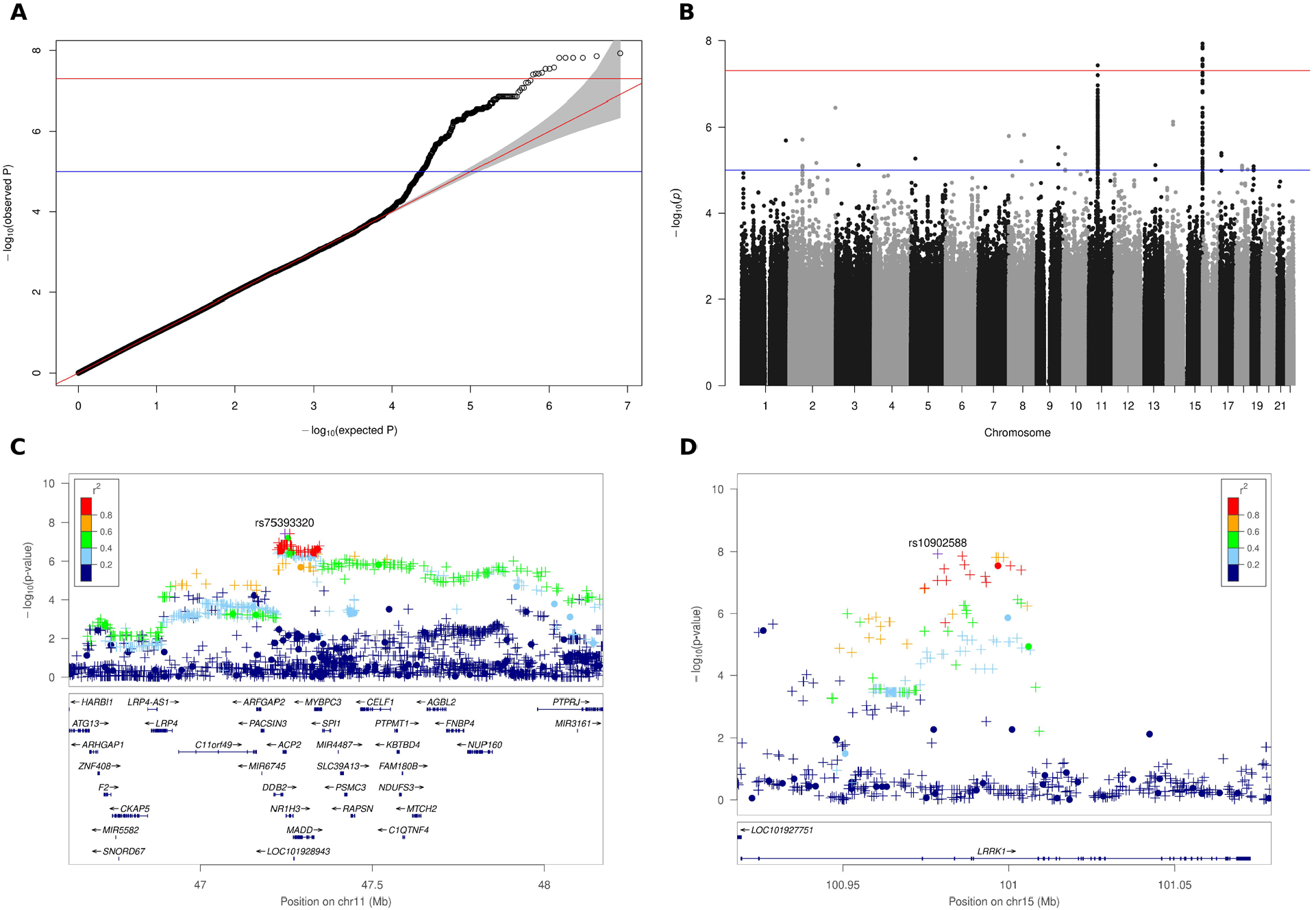
Association plots for anti-NMDAR encephalitis. **A** QQ plot of association analysis for 8,073,349 variants. The plot shows deviation from the null distribution in the upper tail, which corresponds to variants with the strongest evidence for association. **B** Manhattan plot of the association results. The plot shows –log10 marker-wise P-values against their genomic base-pair position. The red line indicates the genome-wide significance threshold of 5 × 10^−8^. **C** LocusZoom plot for the association between anti-NMDAR encephalitis and variants on chromosome 11 in the genomic region from 46.6 to 48.2 Mb. A circle represents a genotyped and a plus symbol an imputed variant. The r^2^ metric displays the pairwise LD between the leading and the respective variant. Gene positions are present in the bottom part. **D** LocusZoom plot for associations on chromosome 15 in the genomic region from 100.9 to 101.1 Mb.

**Table 2.**
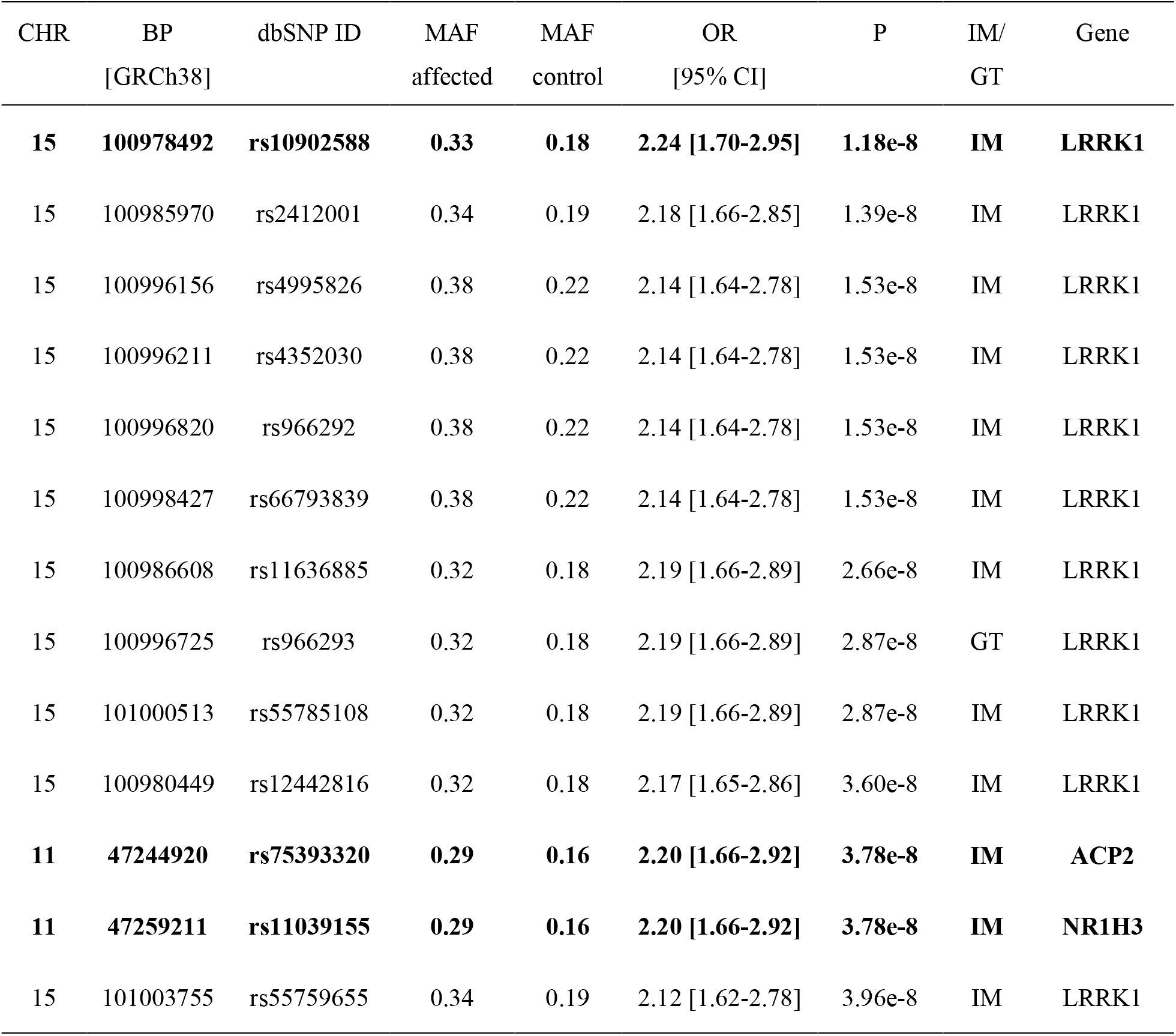
Identified associations with a P-value < 5 × 10^−8^. The top-SNPs at each locus are highlighted in bold, with rs75393320 and rs11039155 on chromosome 11 having the same P-value. IM = imputed, GT = genotyped.

Additional analyses of subpopulations defined by tumor status and age of onset yielded no genome-wide significant associations. In the previous GWAS, we observed a weak association between patients with anti-NMDAR encephalitis and the HLA-B*07:02 allele, preferentially in the patients with late disease onset. We did not confirm this association in the current GWAS. We were not able to analyze the recently reported association with HLA-DRB1*16:02 in a Chinese population^23^ because the frequency of this allele is very low in the German population and no patient in our sample and only one control individual carried this allele. We did not detect any novel significant HLA associations.

As readily apparent in **Figure 1C**, the significant variants on chromosome 11 are located in a gene-rich area with numerous further variants in high-LD with the leading variant and P-values less than 1 × 10^−5^. Coloc analysis showed colocalization with a PP4 > 0.7 between the sum of GWAS variants and GTEx eQTLs for the 3 genes *NR1H3, ACP2*, and MAP Kinase Activating Death Domain (*MADD*) on chromosome 11 in brain tissues and for the four genes *NR1H3, ACP2*, Damage Specific DNA Binding Protein 2 (*DDB2*) and *C11orf49* in various immune cells (**Supplementary Figures S2 and S3**). We did not identify any single variant with a PP4 > 0.7. In contrast, we found no colocalizing eQTL signals for the GWAS signal on chromosome 15.

## Discussion

Except for the HLA complex, the genetic determinants of antibody-mediated encephalitides are unknown. The collection of sufficiently large sample sizes for genetic analyses is hampered by the low disease prevalence of anti-NMDAR encephalitis, which is estimated to be around 0.6/100,000 population.^2^ Despite the small sample size, we were able to find two distinct genomic regions on chromosomes 11 and 15 harboring genome-wide significant disease-associated variants. The locus on chromosome 15 encompasses ∼ 100.000 bp and contains only one gene, *LRRK1*. While genetic variants in the homolog *LRRK2* are the most common cause of autosomal dominant Parkinson’s disease, no neurologic diseases are currently linked to *LRRK1*. In mice, *LRRK1* and *LRRK2* complement each other at least partially in the nervous system, because only deficiency of both proteins causes a neurodegenerative phenotype and both proteins regulate autophagy.^24^ *LRRK1* is expressed in B-cells and monocytes, suggesting a role in the immune system.^25^ Indeed, *LRRK1*-deficient mice show alterations of B-cell development, failure to produce IgG3 class antibodies in response to non-T-cell dependent antigens, and a proliferation and survival defect upon B-cell receptor stimulation.^26^ Yet, there is currently no known connection between *LRRK1* and autoimmunity. However, it is intriguing to speculate, that LRRK1-mediated control of non-T-cell dependent B-cell activation could be dysregulated in patients with anti-NMDAR encephalitis. Indeed, this could explain the observation of frequent non-mutated, germ-line encoded NMDAR antibodies in patients,^27^ the childhood and early adult manifestation, and the coexistence of additional autoantibodies.^28^

The borders of the genomic region on chromosome 11 harboring the second association signal are less defined. The region is much larger, exceeding 1 Mb, and comprises multiple genes. To generate a hypothesis concerning putatively causal genes in this region, we employed colocalization analysis between eQTL data from GTEx for different brain regions as well as immune cell eQTL from the Blueprint project. In brain tissues, we found evidence for colocalization between the genes *ACP2* and *MADD* with eQTL for cerebellum and *NR1H3* with eQTL for the hypothalamus. While it is well known that the hippocampus is a prime target of anti-NMDAR encephalitis, the ubiquitous expression of NMDA receptors containing the GluN1 subunit in the brain, the manifold symptoms of anti-NMDAR encephalitis and pathologic studies suggest an involvement of most if not all brain regions.^29^ Therefore, we think that cerebellum and hypothalamus are valid target regions. In immune cells, we detected colocalization of the genes *NR1H3, ACP2, DDB2*, and *C11orf49* with eQTL in various immune cells including T-lymphocytes. *NR1H3* and *ACP2* show evidence for colocalization in both, brain and immune cells. Unfortunately, B-lymphocytes/plasma cells, the producers of antibodies, are not represented in the Blueprint data. Of the genes identified in the colocalization analysis *NR1H3* encoding the liver x receptor alpha (LXRα) is the best functional candidate. LXRα is a transcription factor whose activation inhibits inflammatory processes.^30^ In the CNS LXRα agonists inhibit proinflammatory cytokine production by microglia and astrocytes.^31^ Knockout of LXRα in brain endothelial cells led to blood-brain-barrier dysfunction, inflammation, and increased transendothelial mononuclear cell migration.^32^ ACP2 is a lysosomal acid phosphatase employed in lysosomal protein degradation, MADD is an adaptor protein involved in transmitting tumor necrosis factor alpha (TNF-α) induced apoptotic signals, DDB2 is involved in DNA repair, e.g. after ultraviolet light damage, and C11orf49 encodes a protein of unknown function.

In contrast to our first GWAS of antibody-mediated encephalitis, we identified two independent genome-wide significant associations in this study. There are three important differences between our previous GWAS and the current one. First, doubling of the sample size led to larger statistical power; second, we carefully removed population outliers; and third, we chose a different set of covariates in the logistic regression model. In contrast to the first GWAS, we included only sex and the first PC in the current analysis. Scree plot analysis suggested using only one PC which might in part be due to the stringent exclusion of ethnic outliers and careful control-matching. Another difference to the first GWAS is the exclusion of age as a covariate.

Genetic variants are stable throughout life. For common late-onset diseases significantly younger controls than patients warrant inclusion of age as a covariate since many controls will still develop the disease during their lifetime. However, in this study the disease is rare and the controls are significantly older than the patients. Including age as a covariate led to partial masking of the signals contributing to the effects in this GWAS. The major shortcoming of this study is its small sample size, which on the one hand limits the power to detect true variant-disease associations and on the other hand did not allow to include an independent replication sample thereby increasing the likelihood of false positives. In summary, we performed a GWAS of anti-NMDA receptor encephalitis and identified two independent genome-wide significant association signals. Both regions contain putative functional candidate genes and eQTL for three genes show significant colocalization with the association signal on chromosome 11.

## Supporting information

STREGA Checklist

## Data Availability

Summary level genetic data for all variants with P-values < 1e-04 are available from the corresponding author on reasonable request to any qualified investigator.

## Acknowledgment

The work was supported by members of the GENERATE network, who contributed to patient recruitment, data acquisition and entry. All members of the GENERATE network as of March 2021 are indicated in **Supplementary Table S1**.

Patient recruitment in the Czech Republic was supported by the Charles University project GA UK No 746120.

Petr Marusic (Department of Neurology, Charles University, Second Faculty of Medicine and Motol University Hospital, Prague, Czech Republic) helped with patient selection and clinical data collection for the Czech participants.

The popgen 2.0 network (P2N; controls) is supported by the Medical Faculty of the University of Kiel.

The Genotype-Tissue Expression (GTEx) Project was supported by the Common Fund of the Office of the Director of the National Institutes of Health, and by NCI, NHGRI, NHLBI, NIDA, NIMH, and NINDS. The data used for the analyses described in this manuscript were obtained from: dbGaP accession number phs000424.v7.p2.

