## Supplementary Table S1 for "Genome-wide association study identifies two new loci associated with anti-NMDAR encephalitis"

| Name | Affiliation |
|---|---|
| Markus Kraemer | Alfried Kupp Krankenhaus Rüttenscheid, Department of Neurology, Essen, Germany |
| Wolfgang Heide | Allgemeines Krankenhaus Celle, Department of Neurology, Celle, Germany |
| Stephan Schreiber | Asklepios Fachkliniken Brandenburg, Department of Neurology, Berlin, Germany |
| Marina Entscheva | Asklepios Fachklinikum Lübben, Department of Neurology, Lübben, Germany |
| Jürgen Hartmut Faiss | Asklepios Fachklinikum Teupitz, Department of Neurology, Teupitz, Germany |
| Robert Berger | Asklepios Klinik Hamburg Altona, Department of Neurology, Hamburg, Germany |
| Oliver Stammel | Asklepios Klinik Hamburg Barmbek, Department of Neurology, Hamburg, Germany |
| Anna Hoffmann | Asklepios Klinik Harburg, Department of Neurology, Hamburg, Germany |
| Günter Seidel | Asklepios Klinik Nord, Department of Neurology, Hamburg, Germany |
| Robert Handreka | Carl-Thiem-Klinikum Cottbus, Department of Neurology, Cottbus, Germany |
| Harald Prüß | Charité Berlin, Department of Neurology and Experimental Neurology, Berlin, Germany |
| Josef Priller | Charité Berlin, Department of Psychology and Psychiatry, Berlin, Germany |
| Carsten Finke | Charité Berlin/NeuroCure, Department of Neurology, Berlin, Germany |
| Paul Friedemann | Charité Berlin/NeuroCure, Department of Neurology, Berlin, Germany |
| Peter Körtvélyessy | Charité Berlin/NeuroCure, Department of Neurology, Berlin, Germany |
| Henning Stolze | Diako Krankenhaus Flensburg, Department of Neurology, Flensburg, Germany |
| Astrid Blaschek | Dr. von Haunersches Kinderspital, Department of Neuropediatrics, München, Germany |
| Sebastian Bauer | Epilepsy Center Frankfurt Rhine-Main and Department of Neurology, Unversity Hospital Frankfurt, Frankfurt am Main, Germany |
| Felix Rosenow | Epilepsy Center Frankfurt Rhine-Main and Department of Neurology, Unversity Hospital Frankfurt, Frankfurt am Main, Germany |
| Kai Siebenbrodt | Epilepsy Center Frankfurt Rhine-Main and Department of Neurology, Unversity Hospital Frankfurt, Frankfurt am Main, Germany |
| Susanne Knake | Epilepsy Center Hessen, University Hospital Marburg, Marburg, Germany |
| Benjamin Wunderlich | Ernst-von-Bergmann-Klinikum, Department of Neurology, Berlin, Germany |
| Sven Ehrlich | Fachkrankhaus Hubertusburg GmbH, Department of Neurology and Neurological Intensive Care, Wermsdorf, Germany |
| Lena Edelhoff | Kath. Marienkrankenhaus gGmbH, Department of Neurology, Hamburg, Germany |
| Judith Wagner | Keple University Hospital, Department of Neurology, Linz, Austria |
| George Trendelenburg | KKH Freiberg, Department of Neurology, Freiberg, Germany |

GENERATE | German Network for Research on Autoimmune Encephalitis

| Name | Affiliation |
|---|---|
| Anna Gorsler | Kliniken Beelitz GmbH, Department of Neurology, Beelitz-Heilstätten, Germany |
| Sebastian Baatz | Klinikum Altenburger Land GmbH, Department of Neurology, Altenburg, Germany |
| Sonka Benesch | Klinikum Aschaffenburg-Alzenau, Department of Neurology, Aschaffenburg, Germany |
| Matthias von Mering | Klinikum Bremen-Nord, Department of Neurology, Bremen, Germany |
| Armin Grau | Klinikum der Stadt Ludwigshafen am Rhein, Department of Neurology, Dossenhaim, Germany |
| Christian Urbanek | Klinikum der Stadt Ludwigshafen am Rhein, Department of Neurology, Dossenhaim, Germany |
| Gernot Reimann | Klinikum Dortmund, Department of Neurology, Dortmund, Germany |
| Tobias Neumann-Haefelin | Klinikum Fulda, Department of Neurology, Fulda, Germany |
| Thomas Pfefferkorn | Klinikum Ingolstadt GmbH, Department of Neurology, Ingolstadt, Germany |
| Sascha Berning | Klinikum Osnabrück, Department of Neurology, Osnabrück, Germany |
| Christoph Kellinghaus | Klinikum Osnabrück, Department of Neurology, Osnabrück, Germany |
| Michael Nagel | Klinikum Osnabrück, Department of Neurology, Osnabrück, Germany |
| Andreas Binder | Klinikum Saarbrücken gGmbH, Department of Neurology, Saarbrücken, Germany |
| Mona Dreeesmann | Klinikum Westbrandenburg, Department of Neuropediatrics and Social Pediatrics, Potsdam, Germany |
| Fatme Seval Ismail | Knappschaftskrankenhaus Bochum, Department of Neurology, Bochum, Germany |
| Ulrich Hofstadt-van Oy | Knappschaftskrankenhaus Dortmund-Klinikum Westphalen, Department of Neurology, Dortmund, Germany |
| Christian Bien | Krankenhaus Mara, Epilepsy Centre Bethel, Bielefeld, Germany |
| Marcel Gebhard | Krankenhaus Martha-Maria Halle, Department of Neurology, Halle (Saale), Germany |
| Frank Hoffmann | Krankenhaus Martha-Maria Halle, Department of Neurology, Halle (Saale), Germany |
| Andrea Kraft | Krankenhaus Martha-Maria Halle, Department of Neurology, Halle (Saale), Germany |
| Franz Blaes | Kreiskrankenhaus Gummersbach, Department of Neurology, Gummersbach, Germany |
| Corinna Bien | Labor Krone, Bad Salzuflen, Germany |
| Andeas Linsa | Lausitzer Seeland Klinikum GmbH, Department of Neurology, Hoyerswerda, Germany |
| Katharina Eisenhut | LMU München, Institute of Clinical Neuroimmunology, München, Germany |
| Joachim Havla | LMU München, Institute of Clinical Neuroimmunology, München, Germany |
| Franziska Thaler | LMU München, Institute of Clinical Neuroimmunology, München, Germany |

**GENERATE** German Network for Research on Autoimmune Encephalitis

| Name | Affiliation |
|---|---|
| Tanja Kümpfel | LMU München, Klinikum Großhadern, Institute of Clinical Neuroimmunology, München, Germany |
| Til Menge | LVR-Klinikum Düsseldorf, Department of Neurology, Düsseldorf, Germany |
| Manuel Friese | Medical Center Hamburg-Eppendorf, Department of Neurology and Institute for Neuroimmunology and Multiple Sclerosis, Hamburg, Germany |
| Max Kaufmann | Medical Center Hamburg-Eppendorf, Department of Neurology and Institute for Neuroimmunology and Multiple Sclerosis, Hamburg, Germany |
| Martin Stangel | MHH Hannover, Department of Neurology, Hannover, Germany |
| Kurt-Wolfram Sühs | MHH Hannover, Department of Neurology, Hannover, Germany |
| Corinna Trebst | MHH Hannover, Department of Neurology, Hannover, Germany |
| Jost Obrocki | Regio-Klinik Elmshorn, Department of Psychiatry, Elmshorn, Germany |
| Jens Schaumberg | Sana Kliniken Lübeck, Department of Neurology, Lübeck, Germany |
| Ilya Ayzenberg | St. Josef-Hospital, Ruhr-University Bochum, Department of Neurology, Bochum, Germany |
| Kerstin Hellwig | St. Josef-Hospital, Ruhr-University Bochum, Department of Neurology, Bochum, Germany |
| Christos Krogias | St. Josef-Hospital, Ruhr-University Bochum, Department of Neurology, Bochum, Germany |
| Friedrich Ebinger | St. Vincenz-Krankenhaus Paderborn, Department of Neuropediatrics St. Louise, Paderborn, Germany |
| Alexander Finke | Städtisches Klinikum Lüneburg, Department of Neurology, Lüneburg, Germany |
| Marie-Luise Mono | Stadtspital Triemli, Department of Neurology, Zürich, Swiss |
| Daniel Bittner | Südklinikum Nordhausen, Department of Neurology, Nordhausen, Germany |
| Stefan Bittner | Universitätsmedizin Mainz, Department of Neurology, Mainz, Germany |
| Simone Tauber | University Hospital Aachen, Department of Neurology, Aachen, Germany |
| Martin Häusler | University Hospital Aachen, Department of Neuropediatrics and Social Pediatrics, Aachen, Germany |
| Annette Baumgartner | University Hospital Basel, Department of Neurology, Basel, Swiss |
| Anne-Katrin Pröbstel | University Hospital Basel, Department of Neurology, Basel, Swiss |
| Stephan Rüegg | University Hospital Basel, Department of Neurology, Basel, Swiss |
| Sarah Bernsen | University Hospital Bonn, Department of Neurodegenerative Diseases and Gorontopsychiatry, Bonn, Germany |
| Alexandra Philipsen | University Hospital Bonn, Department of Psychiatry and Psychotherapy, Bonn, Germany |
| Hendrik Rohner | University Hospital Bonn, Department of Psychiatry and Psychotherapy, Bonn, Germany |
| Philip Hillebrand | University Hospital Bonn, Pediatric Clinic, Bonn, Germany |
| Sigrid Wöpking | University Hospital Dresden, Department of Neurology, Dresden, Germany |

GENERATE | German Network for Research on Autoimmune Encephalitis

| Name | Affiliation |
|---|---|
| Michael Karenfort | University Hospital Düsseldorf, Department of General Pediatrics, Neonatology and Pediatric Cardiology, Center for Social Pediatrics (SPZ)/Neuropediatrics, Düsseldorf, Germany |
| Nico Melzer | University Hospital Düsseldorf, Department of Neurology, Düsseldorf, Germany |
| Sven Meuth | University Hospital Düsseldorf, Department of Neurology, Düsseldorf, Germany |
| Saskia Jania Räuber | University Hospital Düsseldorf, Department of Neurology, Düsseldorf, Germany |
| Marius Ringelstein | University Hospital Düsseldorf, Department of Neurology, Düsseldorf, Germany |
| Regina Trollmann | University Hospital Erlangen, Department of Neuropediatrics, Erlangen, Germany |
| Raphael Reinecke | University Hospital Frankfurt am Main, Department of Neurology, Frankfurt am Main, Germany |
| Dominique Endres | University Hospital Freiburg, Department of Psychiatry and Psychotherapy, Freiburg, Germany |
| Karin Storm van`s Gravesande | University Hospital Freiburg, Pediatric Clinic, Freiburg, Germany |
| Felix von Poderwils | University Hospital Greifswald, Department of Neurology, Greifswald, Germany |
| Steffen Syrbe | University Hospital Heidelberg, Angelika-Lautenschläger-Klinik, Devision of Pediatric Epileptology, Centre for Pediatrics and Asolescent Medicine, Heidelberg, Germany |
| Bettina Balint | University Hospital Heidelberg, Department of Neurology, Heidelberg, Germany |
| Brigitte Wildemann | University Hospital Heidelberg, Department of Neurology, Heidelberg, Germany |
| Carolin Baade-Büttner | University Hospital Jena, Department of Neurology, Jena, Germany |
| Christian Geis | University Hospital Jena, Department of Neurology, Jena, Germany |
| Chung Ha-Yeun | University Hospital Jena, Department of Neurology, Jena, Germany |
| Jonathan Wickel | University Hospital Jena, Department of Neurology, Jena, Germany |
| Michael Malter | University Hospital Köln, Department of Neurology, Köln, Germany |
| Walid Fazeli | University Hospital Köln, Department of Neuropediatrics, Köln, Germany |
| Muriel Stoppe | University Hospital Leipzig, Department of Neurology, Leipzig, Germany |
| Florian Then Bergh | University Hospital Leipzig, Department of Neurology, Leipzig, Germany |
| Johannes Piepgras | University Hospital Mainz, Department of Neurology, Mainz, Germany |
| Valentin Held | University Hospital Mannheim, Department of Neurology, Mannheim, Germany |
| Lara Zieger | University Hospital Marburg, Department of Neurology, Marburg, Germany |
| Marco Gallus | University Hospital Münster, Department of Neurology and Institute of Translational Neurology, Münster, Germany |

GENERATE | German Network for Research on Autoimmune Encephalitis

| Name | Affiliation |
|---|---|
| Oliver Grauer | University Hospital Münster, Department of Neurology and Institute of Translational Neurology, Münster, Germany |
| Stjepana Kovac | University Hospital Münster, Department of Neurology and Institute of Translational Neurology, Münster, Germany |
| Christoph Lehric | University Hospital Münster, Department of Neurology and Institute of Translational Neurology, Münster, Germany |
| Jan Lünemann | University Hospital Münster, Department of Neurology and Institute of Translational Neurology, Münster, Germany |
| Gerd Meyer zu Hörste | University Hospital Münster, Department of Neurology and Institute of Translational Neurology, Münster, Germany |
| Heinz Wiendl | University Hospital Münster, Department of Neurology and Institute of Translational Neurology, Münster, Germany |
| Andre Dik | University Hospital Münster, Department of Neurology, Münster, Germany |
| Catharina Groß | University Hospital Münster, Department of Neurology, Münster, Germany |
| Kristin Stefanie Melzer | University Hospital Münster, Department of Neurology, Münster, Germany |
| Johanna Maria Helena Rau | University Hospital Münster, Department of Neurology, Münster, Germany |
| Christine Strippel | University Hospital Münster, Department of Neurology, Münster, Germany |
| Constanze Mönig | University Hospital Münster, Institute of Translational Neurology, Münster, Germany |
| Karsten Witt | University Hospital of Neurology, Oldenburg, Germany |
| Robert Weissert | University Hospital Regensburg, Department of Neurology, Nittendorf, Germany |
| Marina Flotats-Bastardas | University Hospital Saarland, Department of Neuropediatrics, Homburg, Germany |
| Simon Schuster | University Hospital Schleswig Holstein, Center for Integrative Psychiatry, Lübeck, Germany |
| Ina Schröder | University Hospital Schleswig Holstein, Department of Neurology and Institute of Clinical Chemistry, Kiel, Germany |
| Frank Leypoldt | University Hospital Schleswig Holstein, Department of Neurology and Institute of Clinical Chemistry, Kiel/Lübeck, Germany |
| Andreas van Baalen | University Hospital Schleswig Holstein, Department of Neuropediatrics, Kiel, Germany |
| Justina Dargvainiene | University Hospital Schleswig Holstein, Institute of Clinical Chemistry, Kiel, Germany |
| Martina Jansen | University Hospital Schleswig Holstein, Institute of Clinical Chemistry, Kiel, Germany |
| Ina-Isabelle Schmütz | University Hospital Schleswig Holstein, Institute of Clinical Chemistry, Kiel, Germany |
| Gesa Schreyer | University Hospital Schleswig Holstein, Institute of Clinical Chemistry, Kiel, Germany |
| Klaus-Peter Wandinger | University Hospital Schleswig Holstein, Institute of Clinical Chemistry, Kiel/Lübeck, Germany |

GENERATE | German Network for Research on Autoimmune Encephalitis

| Name | Affiliation |
|---|---|
| Halime Gül | University Hospital Ulm, Department of Neurology, Ulm, Germany |
| Jan Lewerenz | University Hospital Ulm, Department of Neurology, Ulm, Germany |
| Loana Penner | University Hospital Ulm, Department of Neurology, Ulm, Germany |
| Makbule Senel | University Hospital Ulm, Department of Neurology, Ulm, Germany |
| Hayrettin Tumani | University Hospital Ulm, Department of Neurology, Ulm, Germany |
| Methab Türedi | University Hospital Ulm, Department of Neurology, Ulm, Germany |
| Luise Appeltshauser | University Hospital Würzburg, Department of Neurology, Würzburg, Germany |
| Kathrin Doppler | University Hospital Würzburg, Department of Neurology, Würzburg, Germany |
| Claudia Sommer | University Hospital Würzburg, Department of Neurology, Würzburg, Germany |
| Dirk Fitzner | University Medical Center Göttingen, Department of Neurology, Göttingen, Germany |
| Jens Schmidt | University Medical Center Göttingen, Department of Neurology, Göttingen, Germany |
| Niels Hansen | University Medical Center Göttingen, Department of Psychiatry, Göttingen, Germany |
| Kevin Rostasy | Vestische Kinder- und Jugendklinik Datteln, Department of Neuropediatrics, Datteln, Germany |
| Michael Adelmann | Vitos Weil-Lahn, Department of Neurology, Usingen, Germany |

**Supplementary Table S1. Members of the GENERATE network (March 2021)**
