## Supplementary Table S2 for "Genome-wide association study identifies two new loci associated with anti-NMDAR encephalitis"

| | Cases | Controls | Total |
|---|---|---|---|
| Samples started with | 212 | 1,219 | 1,431 |
| | | **Number of excluded samples** | |
| Genotyping not successfull | 0 | NA | 0 |
| Sample call rate < 85% | 3 | 0 | 3 |
| Sample Call rate < 98% or heterozygosity Rate > 3xSD | 13 | 3 | 16 |
| Cryptic relatedness > third degree relatives | 5 | 13 | 18 |
| Missing or discrepant sex | 5 | 0 | 5 |
| Population Outlier | 8 | 0 | 8 |
| Control matching | - | 613 | 613 |
| **Total samples analyzed** | **178** | **590** | **768** |

**Supplementary Table S2. Sample-based quality control**

Summary of sample-based quality control steps of genotyped data. 34 patients with anti-NMDAR encephalitis and 629 healthy controls had to be excluded based on quality control procedures and control matching.
