## Supplementary Figures for "Genome-wide association study identifies two new loci associated with anti-NMDAR encephalitis"

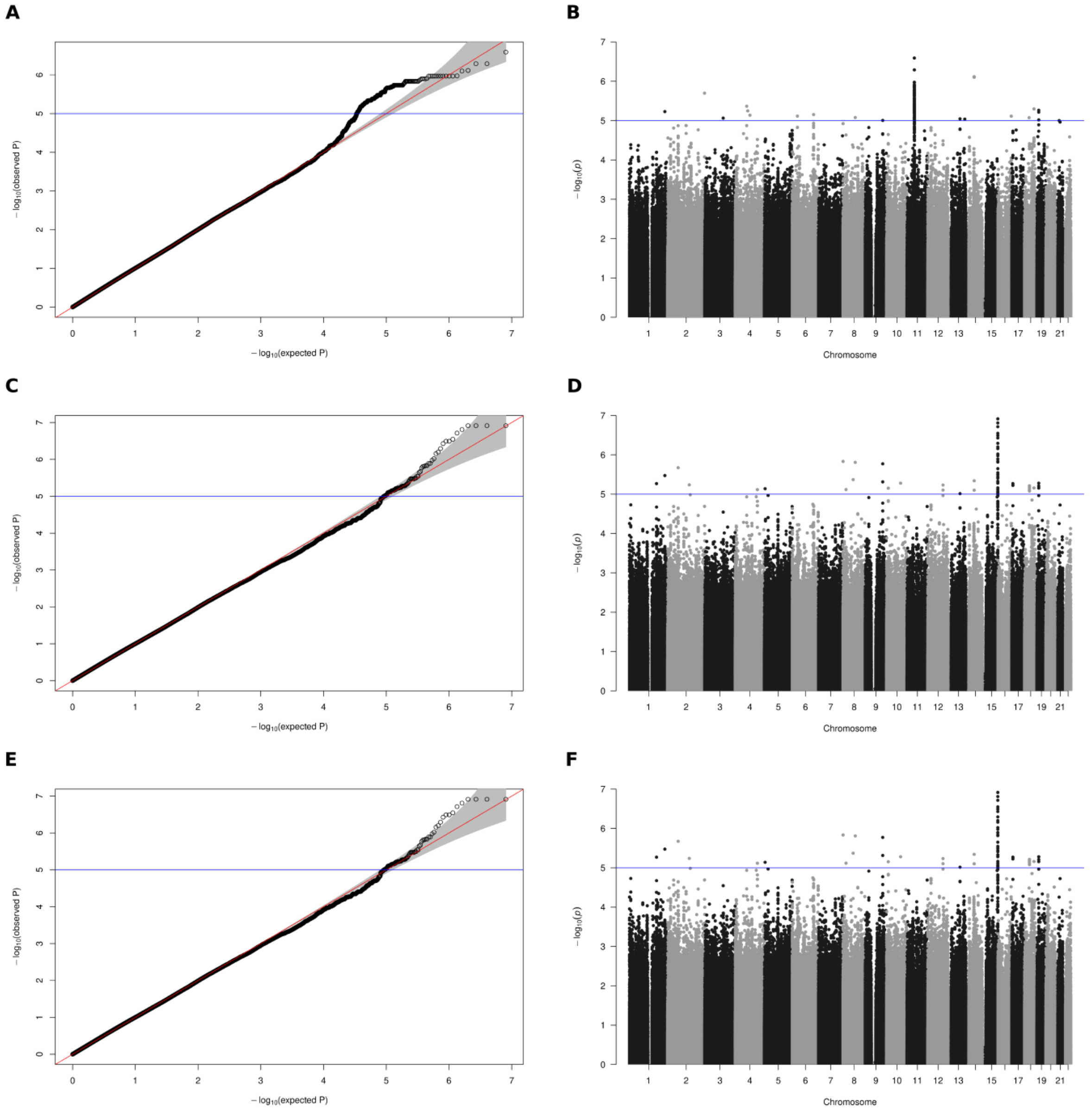


**Supplementary Figure S1**. **Association plots for conditional analyses**

QQ plots (left plot panel) and manhattan plots (right plot panel) for conditional analysis including respectively one of the top SNPs as additional covariate. Analyses included sex, the first PC and rs10902588 (Chr. 15) in panel **A** and **B**, rs75393320 (Chr. 11) in panel **C** and **D** and rs11039155 (Chr. 11) in panel **E** and **F**. The comparison to the original QQ plot (Figure 1A) indicates that the visible deviation from the distribution of P-values under the null hypothesis of no association is mainly due to the top SNPs identified in this GWAS. The manhattan plots confirm that no significant secondary signals are present (only shown for the SNPs with lowest P-values at each locus)

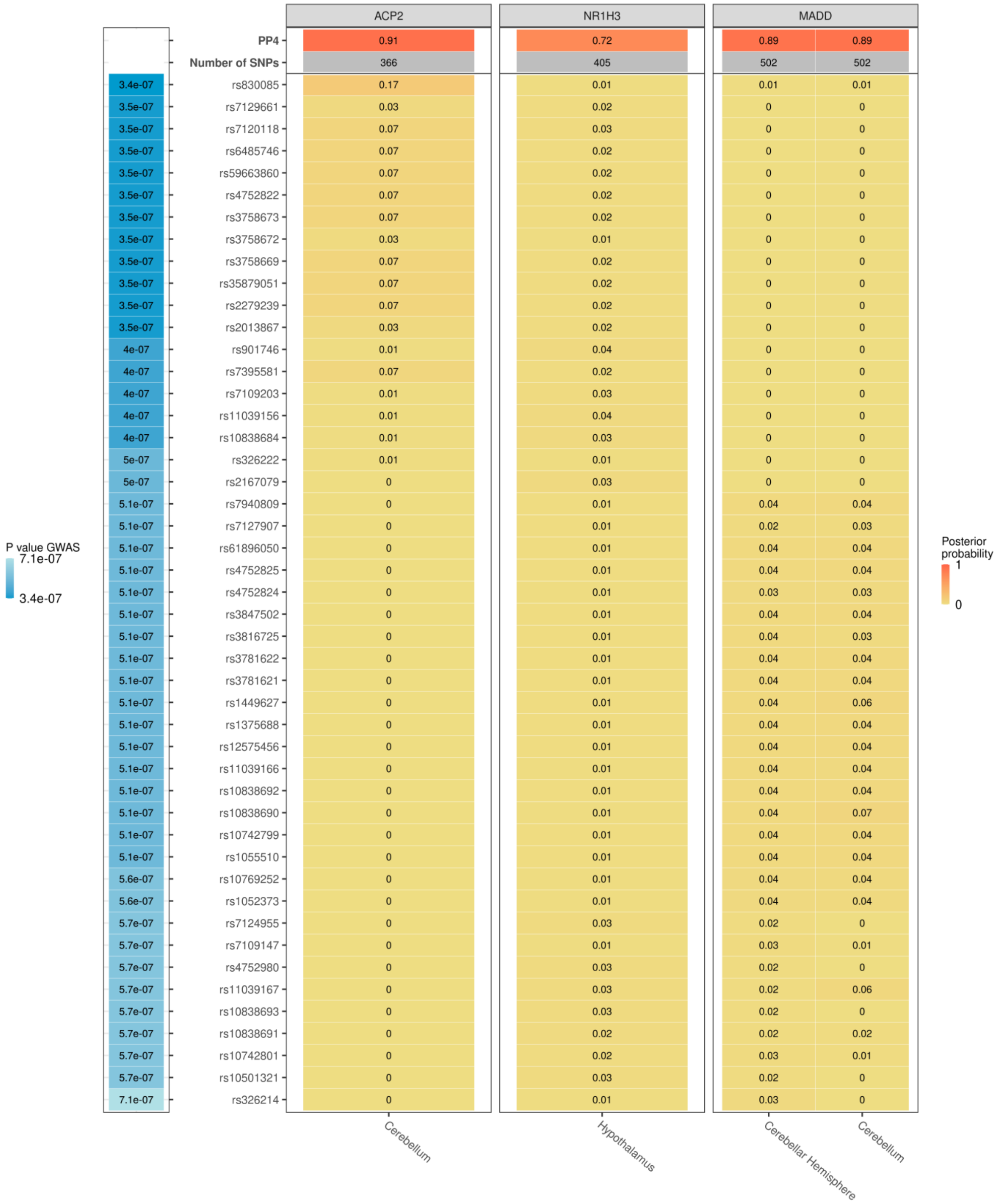


**Supplementary Figure S2. Colocalization results for brain tissues**

Gene- and SNP-wise results of the colocalization analysis for brain tissues represented in GTEx types. Only genes with a PP4 > 0.7 and variants with a P-value < $10^{-5}$ are shown.

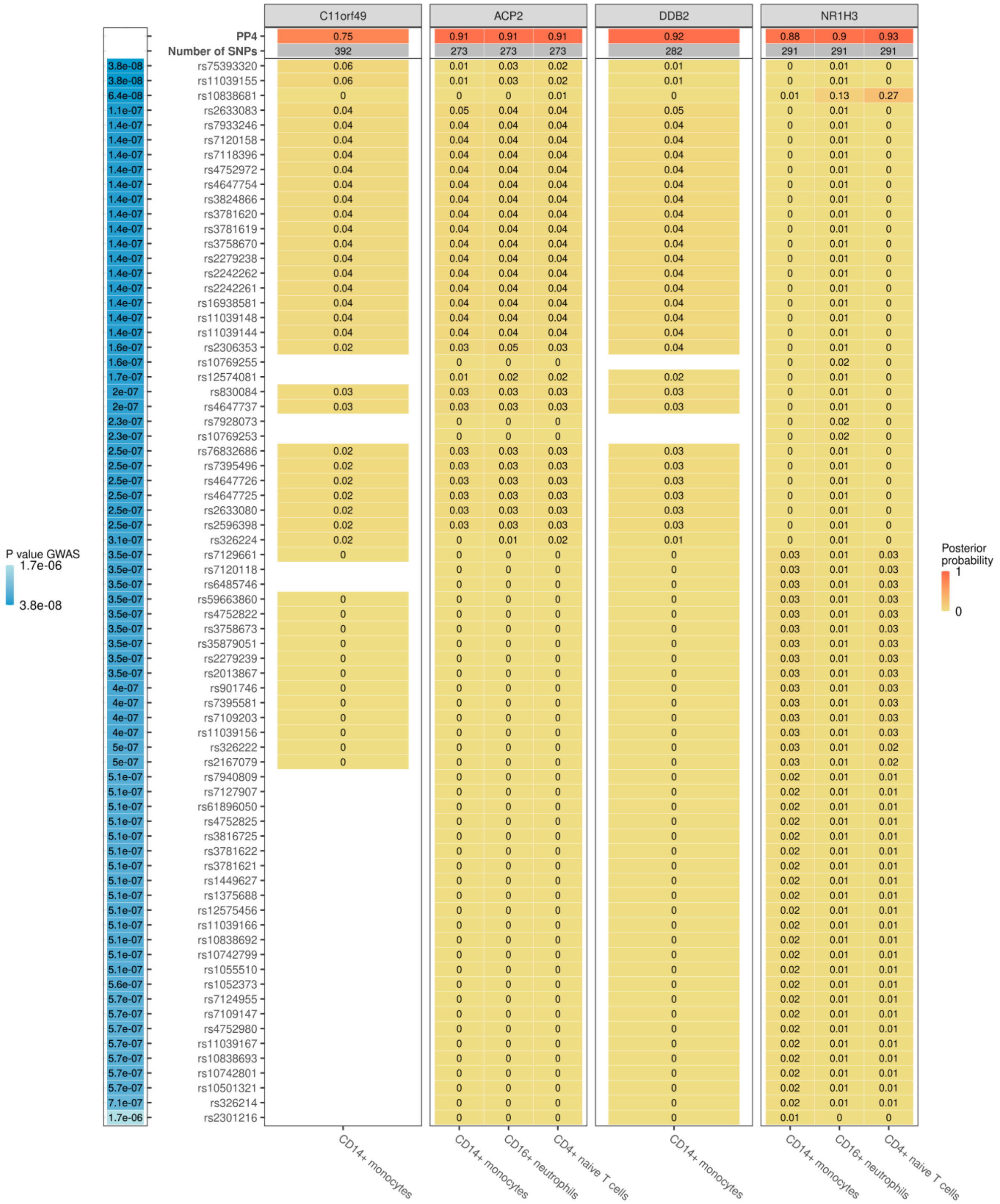


**Supplementary Figure S3**. **Colocalization results for immune cells**

Gene and SNP-wise results of the colocalization for immune cells represented in the Blueprint dataset. Only genes with a PP4 > 0.7 and variants with a P-value < $10^{-5}$ are shown.
